# A Scoping Review on Influence of Socioeconomic Status on Antenatal Care Utilization and Pregnancy Outcomes in Sub-Saharan Africa

**DOI:** 10.1101/2024.01.11.24301063

**Authors:** Adeyemo Queen Esther, Haphsheitu Yahaya, Ajayi O. Esther, Priscilla Aboagye-Mensah, Adeyemo J. Blessing, Theckla E. Ikome

## Abstract

Maternal and perinatal mortality are the most adverse pregnancy outcomes of public health concerns. Although, slowly declining, Sub-Saharan Africa (SSA), has been reported as one of the regions with the highest incidence globally. Regions in SSA where these problems are prominent have been reported to have poor access to antenatal care services. Since socioeconomic factors are major factors influencing the use of antenatal care services and pregnancy outcomes. This study, therefore, aimed to explore the socioeconomic determinants of antenatal care utilization and pregnancy outcomes in Sub-Saharan countries. Studies were systematically searched using credible search engines, whereby 82 studies based on the selection criteria from eight countries with reported maximum burden of study were found. Consistently across all reviewed studies, poor socioeconomic status was a significant determinant of Antenatal care utilization thus leading to poor pregnancy outcomes, particularly, low income, and education. The impact of occupation on the other hand has been poorly studied. Poor socioeconomic factors also limit the use of antenatal care services, increasing the burden of the deaths. The study therefore submits that, interventions, and policies to reduce maternal and perinatal mortality should focus on improving pregnant women’s’ lives by improving access to antenatal care services pre- and postnatal period.

## Introduction

The prenatal and postpartum periods of pregnancy are crucial for ensuring smooth and long-lasting transitions to long-term preventive health care because they are a window into future health [1]. Any complications during the pregnancy process, results in adverse pregnancy outcomes. Adverse pregnancy outcomes have been a significant public health issue around the world. These outcomes play a major role in maternal and perinatal mortality rates worldwide, with women of African descent more susceptible than those in developed countries [2].

Nearly 800 women die from pregnancy related causes. This is despite the claim that, the global maternal mortality rate (MMR) decreased by 38% between 2000 and 2017. Most maternal deaths occur in developing countries at a rate of 1 in 4,900 compared with 1 in 180 in developed countries, with 75% caused by pregnancy and childbirth-related problems according to WHO report. In the WHO’s African Region, 45 out of 47 countries reported 147,741 maternal deaths in 2010. Shockingly, this situation remains largely unchanged, with 99% of maternal deaths still occurring in underdeveloped nations, and more than half of these deaths happening in sub-Saharan Africa [3].

Perinatal mortality is characterized by severe adverse outcomes for children during pregnancy, and it is also a significant public health concern. Perinatal mortality is deaths occurring in vitro or intrauterine following the completion of 24 weeks of gestation (stillbirth) or within seven days of delivery (early neonatal death) [4]. Perinatal mortality falls into two categories: stillbirths (expressed per 1000 total births) and early neonatal mortality (expressed per 1000 live births). To combat child mortality therefore, Sustainable Development Goal (SDG) 3.2 aims to end preventable deaths of newborns and under-5 children, specifically reducing neonatal mortality to below 12 per 1000 live births and under-5 mortality to less than 25 per 1000 live births by 2030. However, a report by the US National Statistic for the UN SDG revealed that many countries were not on track to meet these targets, especially in low and middle-income countries with unreliable mortality data.

One of the interventions in most African countries to improve pregnancy outcomes is ensuring free antenatal care services [5,6]. Recommendation on reducing adverse pregnancy outcomes has been centered on accelerating healthcare services to pregnant women before, during, and after pregnancy. The use of antenatal care services significantly reduces maternal and perinatal mortality [7]. However, majority of women of reproductive age group does not access antenatal care services or does partially, this utilization is believed to be influenced greatly by the women’s socioeconomic characteristics [8]. Socioeconomic factors such as income level, education and type of occupation are important factors that influence pregnancy outcomes [5,9].

The Social Determinants of Health Theory (SDOH) is used as a framework to guide this research, suggesting that future studies and policies should focus on addressing these socioeconomic factors to improve pregnancy outcomes and reduce maternal and child mortality. Several studies [10,11] have reviewed and studied different aspects of SDOH on pregnancy outcomes and have recommended improving healthcare policies and services to improve pregnancy outcomes. However, with the recent change in the global economy and increased inflation and hardship before the global COVID-19 pandemic and worsened by it, there is need to focus on factors that can improve the living conditions of pregnant women. The objective of this study is therefore to systematically examine and synthesize evidence that supports the impact of socioeconomic economic status; poor education, poor income, and occupation, on poor utilization of antenatal care services and adverse pregnancy outcomes; maternal and perinatal mortality, under the following questions:

Research Questions

1. What is the extent of the impact of poor socioeconomic status disaggregated into the impact of education, occupation, and income on maternal and perinatal mortality in SSA countries with high maternal, perinatal mortality and poor ANC utilization.
2. What are the variations in the socioeconomic factors (impact of education, occupation, and income) affecting ANC utilization as well as maternal and perinatal mortality in SSA with low socioeconomic status.
3. To what extent have these socioeconomic barriers (poor education, occupation, and income) influenced access to antenatal care services among pregnant women from low socioeconomic backgrounds with high maternal and perinatal mortality in SSA

*The following elements can be renamed as needed and presented in any order:*

## Methods

### • Selection of Relevant Studies

Studies were searched using Google Scholar, SCOPUS, PubMed, and Elite search engines, majorly from credible journals; MEDLINE/PubMed, Cochrane Library, SCOPUS, CINAHIL, Science Direct, and Directory of Open Access Journal (DOAJ) databases systematically. Specific keywords were grouped into five categories to search for relevant articles across the various databases following strictly PRISMA ScR research guidelines for conducting literature search for scoping reviews. The first category includes terms that describe the type of socioeconomic factors, which includes keywords like “social class”, “Income level”, “Wealth level”, “Education level”, “Occupation level”, “Standard of living”, “Social stratification” and “Economic background”. The second category includes terms that describe maternal and perinatal mortality, such as “maternal death”, “Maternal fatality”, “Pregnancy-related death”, “Obstetric mortality”, “Maternal morbidity”, and “Perinatal death”, “Perinatal fatality”, “Fetal mortality”, “Stillbirth”, “Neonatal mortality”, “Infant mortality”, “Newborn death”, “Preterm birth”, “Antenatal death”, “Intrauterine fetal demise”, “Fetal demise”. The third category covers access and utilization to ANC or Prenatal care services, which includes keywords like “utilization,” “access,” “use,” “barriers,” “facilitators,” and “decisions.” The fourth category represents the determinants, which includes keywords like “factors” and “determinants.” Lastly, the fifth category describes the place of study, which includes “Sub-Saharan Africa”, “Central Africa”, and “Southern Africa”, “East Africa”, “West Africa”. These search terms were combined using the Boolean operators “AND” and “OR.”

### Inclusion Criteria

The search was limited to studies done in Sub-Saharan Africa (SSA) countries with poor socioeconomic status, high maternal mortality, and high perinatal mortality. Sub-Saharan Africa in this paper denotes countries geographically located in the 4 parts of the Sahara (western part, Eastern part, central part, and Southern part of Sub-Sahara Africa). Studies that matched the following requirements were considered for inclusion in the review.

1. Both qualitative and quantitative studies in academic journals published between 2010-2023.
2. Empirical peer-reviewed papers written in English
3. Papers that focused on antenatal care utilization, maternal and perinatal mortality

### Characteristics of the Included Studies

This study was done in the Sub-Saharan African countries with poor level of income, poor literacy rate, poor occupational status, and poor antenatal care utilization yet with high prevalence of maternal and perinatal mortality. After all exclusion criteria were observed, a total of 8 countries were selected out of 49 SSA countries for the study. Three countries from the Eastern Africa; Burundi, South Sudan, and Somalia, 4 from Western Africa; Nigeria, Guinea, Sierra Leone and The Gambia and one country from Northern Africa; Chad. The review involves 82 studies out of the initial 2190 studies conducted between the year 2010 and 2023 after the inclusion criteria was applied (Figure 1). It includes 7 studies done in Burundi, 29 in Nigeria, 8 in Guinea and 8 in The Gambia.

**Figure 1:**
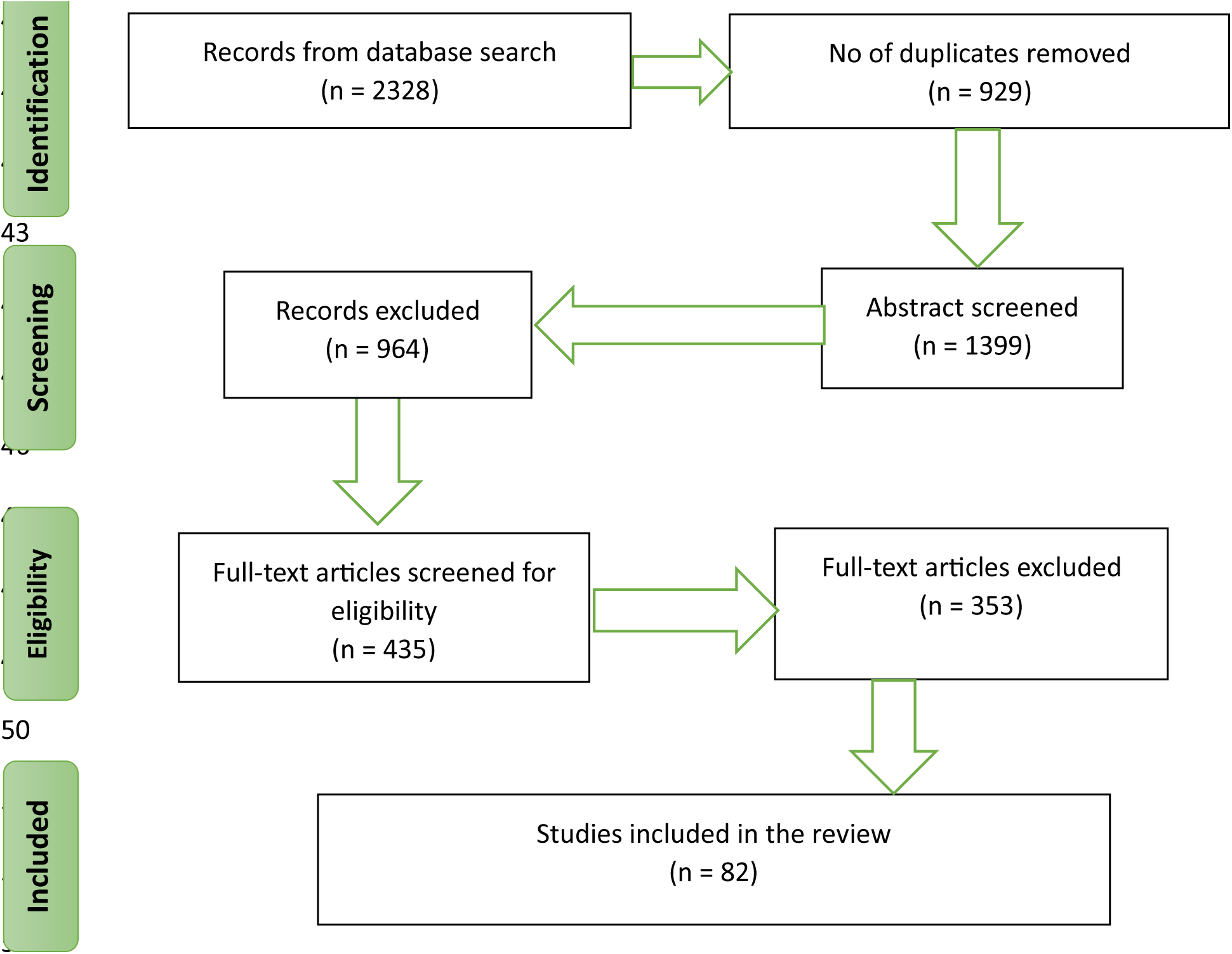
PRISMA Flow chart of the studies included

### Exclusion Criteria

Papers on countries outside Sub-Sahara Africa; papers not related to socioeconomic status in the aspect of low income, high unemployment rate, and low level of education, papers not in the English Language were excluded.

### Outcome Definition

#### Perinatal mortality rate

This is the sum of the number of deaths of a fetus after 28 weeks of viability to before the 7^th^ day of life. Can be divided into stillbirths and early neonatal deaths. (WHO, 2006).

#### Maternal mortality ratio (MMR)

This is defined as the number of maternal deaths during a given period per 100,000 live births during the same period.

Maternal deaths denote the death of a woman during pregnancy or within 42 days of the end of pregnancy from any cause related to pregnancy or aggravated by it and its management irrespective of the duration and site of the pregnancy expressed per 100,000 live births, for a specified period (WHO, 2015)

## Method

## Results

### Impact of Socioeconomic Status (SES) on Maternal Health Care Utilization SES on Maternal Health Care Utilization

Socioeconomic factors like education, income, and occupation were found to significantly impact ANC utilization [12]. Women from higher socioeconomic backgrounds have better access to maternal health services, while those from lower socioeconomic positions report low utilization [1] and poor prognosis [13,14]. Several studies have also found that Socioeconomic barriers (with emphasis on education, occupation, and income) has influenced access to antenatal care services among pregnant women from low socioeconomic backgrounds [15–17]. For instance, a study investigated the availability and quality of emergency obstetric care in The Gambia’s main referral hospital and examined the impact of socioeconomic factors on antenatal care services utilization. The study found that women from low socioeconomic backgrounds were less likely to utilize antenatal care services due to a lack of financial resources and transportation. In addition, the study found that women who lived far away from health facilities had limited access to antenatal care services. The study highlighted the need for policies and programs that address these socioeconomic barriers to improve access and utilization of antenatal care services in The Gambia [18]. Studies have shown that the situation in The Gambia and that of Burundi are similar and as we delve deeper into exploring the impact of the specific socioeconomic status on maternal health care as we progress

### Impact of Education on ANC

The prevalence of Antenatal visits in Burundi is majorly below 6%, only the region of Bujumbura-Mairie has the highest ANC prevalence (24. 6%) and the lowest is in the West region (2.7%) [19]. Despite antenatal care (ANC), delivery and postnatal care being free at the point of use in Burundi, utilization of these services remains low: between 2011 and 2017, only 49% of pregnant women attended at least four ANC visits [20]. Some of the factors identified in this study suggested that factors such as women’s education, their husbands’ education level, where they live, whether they plan to become pregnant, family income, and the household wealth index are factors that influence whether they used antenatal care [19].

Over 46% of maternal deaths in Nigeria occurs among women who did not attend ANC clinics, had little or no education with low wealth status being the predominant factors [21]. Peculiarities exist between the northern part and southern parts of Nigeria. The northern part of Nigeria is particularly known for high burden of maternal mortality. The socioeconomic factors responsible for this disparity in use of care in pregnancy was mostly poor education, coupled with high level of ignorance, cultural dependence, and myths [22,23]. Whereas, in the southern parts of Nigeria, poverty and education were the most predominant factors [21,24,25]. In the rural region of the country, factors such as poor maternal formal education, unemployment, even partners education, household wealth were all significant in this study to affect ANC use [26], whereas in the urban areas there are few variations, though maternal formal education and that of their partners and wealth status were still important factors that influences maternal care usage.

The findings in Guinea are very similar to that of Nigeria. The factors that influence antenatal care utilization was mostly level of education and wealth status. Other factors were cultural in terms of power of decision making, religion and level of media awareness [15,27]. A qualitative study was done to assess the factors affecting the attendance and right timing of antenatal visits in Guinea, the findings were categorised into four factors which were related to accessibility to the care, their attitudes about ANC, knowledge of what it entails and other interpersonal reasons [27].

Jalu et. al claimed that like in other countries reviewed literacy and having formal education played a big role in anti-natal care utilization [28]. Moreover, lack of exposure to health education from health care providers and mass media/radio may also limit the knowledge of the community. This study revealed that an educated mother is more likely to utilize MHC compared to a mother with no education, being consistent with previous studies. Specifically, the odds of ensuring a higher level of utilization are 1.33 times and 2.1 times greater among women with primary and at least secondary education respectively, than uneducated women. The education level of male spouses also has a significant effect on women’s service utilization as women whose partners attained at least primary education are around 1.6 times more likely to benefit from MHC at a higher level [29].

In South Sudan, there is proof that maternal education cannot be overemphasized as long as ANC utilization is concerned. Based on the study conducted in South Sudan, pregnant women with primary and secondary level of education were 2.04 and 2.32 times, respectively, more likely to use ANC than women with no education [30]. This finding agreed with that of [31] that concluded that non-use of ANC services was also significantly higher among illiterate women and odds of non-attendance were lower in urban areas. In Sierre Leone, women with higher level of education were found to have higher odds to access maternal health care than those without formal education [32,33].

### Impact of Income on Maternal Health Care Utilization

A study on Burundi concluded that pregnant women in this country experiences obstacles to timely access and usage of maternal health services due to factors like poverty, distances to medical facilities, and high expenses of health services [20]. Another study conducted in Burundi revealed that there is a highly substantial inverse association between per capita income and maternal mortality [34]. The data suggest that fewer maternal fatalities exist as household income increases, in Burundi. According to this study, there are significant disparities between maternal mortality rates in rich and developing nations. Women in low-income countries are dying because they have poor or no access to healthcare, or because the care they receive is of poor quality [34].

Similarly, income was the biggest identified challenge to ANC visit in Gambia especially in the rural communities where cases of maternal complications were common [18]. The study identified, inability to afford the services, cost of traveling to access the services and lack of means of transportation, all of which relate to influences of income level.

In Nigeria, women from wealthier households are more likely to utilize ANC services than those from poorer households [5,35,36]. In addition to income, the cost of antenatal care was a major barrier to utilization for women in Nigeria [24,37,38]. Interestingly, community level income influences antenatal care utilization. A study conducted in Nigeria assessed the moderation role of community level factors on maternal health care revealed how much this factor can be of influence. The study found that women who reside in poorer and poorly educated communities in Nigeria tend to have less appreciation for using maternal health care services [39]. Studies also found that women with lower income were more likely to delay seeking care due to financial constraints [26,39,40]. Further affirming the importance of income to MHC utilization in Nigeria.

Equally in Guinea the findings were similar to Nigeria, wealth status was a strong determinant for attending ANC. The rich had better access to antenatal care and easier deliveries [15], while majority of the women with low income in Guinea complained of the cost of attending ANC clinics and the ability to afford the care [27].

Somalia has limited and understaffed health care centres that works only a few hours per day, an inadequate payment system of salaries has made it difficult for mothers to pay right bills limiting their access to these antennal services [41]. Another study in Somalia found that women whose monthly household income was below $50 did not attend ANC [42,43]. In Chad an increase in wealth index increases the chances of ANC utilization [30].

The situation in South Sudan is similar to that of Chad whereby enabling resources, such as the household wealth index, were significantly associated with non-use of ANC services [31]. For instance, a study found that women from the top two household wealth quintiles had an increased utilization of the recommended number of four or more ANC visits compared to their counterparts from the lowest two quintiles [43]. Also, women of lower socioeconomic status and who specifically cannot afford to pay the transport fare to the health facility did not utilize ANC.

### Impact of Occupation on use ANC

The sparse studies on occupation as a factor to influence ANC visit in Burundi found out that woman’s likelihood of requesting ANC services from a qualified healthcare expert increases with her occupation, marital status, and money. Limited healthcare infrastructure and a shortage of skilled healthcare workers in rural areas may contribute to poor utilization of ANC services among women living in these areas according to Burundi’s household Survey, 2016.

In The Gambia, a woman’s capacity to access ANC services may depend on her employment condition. In rural Gambia, pregnant women were not given special treatment because they were still expected to perform their household tasks. They had a lot of work to do and few possibilities to take time off for illness. Also, the paucity of resources made it nearly impossible to get prenatal care. Women in polygamous households typically worked in the fields for little pay and received little financial support. Their spouses and other family members did not relieve them of their pregnancy-related domestic duties [43].

In Nigeria, increased employment led to decrease in usage of care, this could be due to the fact that most gainfully employed women in urban areas have limited time to attend ANC clinic due to tight work schedules. The impact of occupation on antenatal care utilization in Nigeria is complex and multifaceted. Women in certain occupations may have more difficulty accessing ANC services due to factors such as long working hours, inadequate maternal leave policies, and lack of flexible work arrangements [26]. On the other hand, women in formal employment, such as civil servants, may have better access to ANC services due to the availability of maternal leave policies and flexible work arrangements that allow them to attend ANC appointments. Additionally, some formal sector employers offer health insurance benefits that cover ANC services, which can increase access and utilization [21].

However, women working in the informal sector, such as traders, may find it challenging to access ANC services due to the nature of their work and the lack of social protections. Several studies have shown that women in low-income occupations, such as agriculture and informal sector workers, have lower utilization of ANC services compared to those in high-income occupations, such as civil servants and professionals [43]. Furthermore, rural women tend to have lower utilization of ANC services compared to urban women, regardless of their occupation [14,44].

In addition to the possible impact of these unhealthy behaviours, being unemployed may have a negative impact on one’s ability to pay for high-quality medical care. The budget on such income, for example, could encourage a limited healthy diet or underuse of healthcare during the vital period of pregnancy, raising the likelihood of an unfavourable outcome if a spouse depends on it to provide nutritious foods and adequate prenatal care [43].

Occupation plays a significant role in determining access to and utilization of ANC services in Nigeria. Women in low-income occupations and those in the informal sector are particularly vulnerable and may require targeted interventions to improve their access to ANC services. Policymakers and stakeholders should prioritize the development of policies and programs that support working women’s access to ANC services, regardless of their occupation, to improve maternal and child health outcomes [35,36,40].

Occupational status of the Guineans was a factor to determine their wealth status. Women who were more empowered or had partners who were, had better ANC attendance as well as better pregnancy outcomes [45,46]. A Somalian study found that 72.0% of the respondents who were housewives did not attended ANC therefore there was a significant statistical association between respondents’ occupation and ANC attendance and there was an association between the occupation of the husband and ANC utilization [42]. In Chad, women had lower odds of non-attendance if their partner worked in agriculture compared to those who had professional jobs [30].

### SES on Pregnancy Outcomes

Several studies have found relationships between SES and pregnancy outcome (maternal and perinatal mortality) in Sub-Saharan Africa. Factors contributing to these high rates of maternal mortality include limited access to maternal healthcare services, low levels of education among women, poverty, and a shortage of skilled birth attendants.

### Impact of Education on Pregnancy Outcomes

Education can influence a woman’s knowledge about healthy pregnancy and childbirth practices, access to healthcare, and decision-making abilities related to their health and that of their baby. Studies have shown that maternal education is positively associated with improved pregnancy outcomes, including reduced risk of stillbirths, preterm birth, and low birth weight. Pregnancy-related problems led to around 303,000 maternal deaths worldwide in 2015 with majority of these deaths occur in developing nations [6]. There is a strong belief that high educational status can improve pregnancy outcomes.

Maternal education is a critical factor that can impact pregnancy outcomes in Burundi. Unfortunately, education is limited in Burundi, particularly for girls, with high rates of illiteracy reported among the population [47]. The average number of ANC visits in Burundi is higher for women who complete tertiary education than for women without any formal education. This may be due to greater knowledge and awareness of the importance of ANC, as well as better access to healthcare services [20]. In Guinea, women who were more educated were more informed about the importance of antenatal care visits and had better access to healthcare too [27,48].

In The Gambia, education has been identified as a crucial factor in reducing both maternal and perinatal mortality rates [49]. Hence, increasing the level of education among women could lead to a significant reduction in maternal mortality in The Gambia [50]. The impact of education on maternal and perinatal mortality is greatly felt in Nigeria, as 42% of maternal mortality was as a result of lack of formal education [21]. Studies such as [5,51,52] further attributed lower rates of stillbirth and other perinatal and maternal mortality in Nigeria to maternal education.

Moving on to Somalia, maternal education was found to be a contributing factor to still births and perinatal mortality among Somali nationals [53]. In a Chad study, it was opined that the socioeconomic level measured through their education (as well as income and occupation) of the mother is an important predictor of malnutrition, poor health and mortality [54]. This affects the food habits and consequently the person ‘s health and may cause mortality of one or both mother and child. A study that was conducted in South Sudan concluded that maternal education was significantly associated with lower risk of unfavorable outcome and that this was primarily with the neo-natal death rate rather than the still birth rate [55]. Finally, a study conducted in Sierre Leone found that children born to educated mothers had higher survival rates compared to those whose mothers were not educated [56].

### Impact of Income on Pregnancy Outcomes

Studies suggest that low-income levels and poverty are significant contributing factors to high mortality rates. Women in low-income households are less likely to access quality antenatal care, skilled birth attendants, and emergency obstetric care. Additionally, poverty affects nutrition, education, and living conditions, all of which can impact maternal and child health outcomes. poverty may limit access to antenatal care, skilled birth attendants, and emergency obstetric care, leading to higher rates of perinatal mortality [15,17,57,58].

A study revealed that there is a significant inverse relationship between per capita income and maternal mortality in Burundi. It reported that as household income increases, maternal deaths decrease, while holding all other factors constant. Additionally, the study also shows a negative relationship between government expenditure on the health sector and overall mortality rate, meaning that an increase in government allocation to health could reduce maternal deaths, but at a lower rate [34].

The situation is not so different In the Gambia, whereby money is a huge barrier to assessing comprehensive care in pregnancy and preventing birth complications to both mother and baby. Consistently, all the studies reviewed on the situation in Gambia, revealed lack of money as a factor leading to high cases of maternal and perinatal mortality in Gambia [18,59,60].

Nigeria, the largest country in terms of economy and population in Africa, has over 60% of her population living slightly above a dollar daily and over 80 million of the population lives in extreme poverty. Majority of the top causes of maternal and perinatal mortality are mostly as a result of poverty, illiteracy and its accompanying ignorance level [61]. Studies also found a strong correlation between poverty and perinatal death with poverty considered to be an underlying factor linked to poor delivery outcomes [9,39]. Categorising these regions into two communities of rural and urban, overall, mortalities are higher in the rural communities where there is marked socioeconomic influence, which majorly is related to poor income, illiteracy and poor gainful employment [51,62,63].

In Somalia, absence of transport fare and low income affected access to proper care [64] and the chances of still births was found to be high among Somali women as a result of low income [53]. Furthermore, Somalia’s GDP per capita was found to affect the rate of maternal mortality [65]. Findings from South Sudan revealed that family income significantly influences infant mortality since it is one of the most important determinants of the standard of living, economic and social welfare [66]. About 50.3% of the respondents were classified as low-income group; this definitely will affect the accessibility of the medical services. Contrary to what was found in other countries, mother’s working status did not influence infant mortality in Sierre Leone [56]. The mother’s working status determines higher income possibilities.

### Impact of Occupation on Pregnancy Outcomes

Occupation is another factor that have an impact on pregnancy outcomes, most women affected by birth complications are usually not skilfully employed as it can affect a woman’s access to healthcare, exposure to harmful substances, and physical demands during pregnancy [60]. In general, occupations that involve exposure to harmful substances, such as agriculture or mining, can increase the risk of adverse pregnancy outcomes, including stillbirths, preterm birth, and low birth weight [67] Women in these occupations may also have limited access to healthcare and face additional physical demands during pregnancy, which can further increase the risk of complications. On the other hand, occupations that provide better access to healthcare and allow for more flexibility, such as office-based jobs, may lead to better pregnancy outcomes. Women in these occupations may be better able to attend prenatal care appointments, manage their workload and stress levels during pregnancy, and take time off if needed [68].

Burundi is one of the poorest countries in the world, with a low socioeconomic status. The majority of the population lives in rural areas, and subsistence agriculture is the primary source of livelihood for many households. The country’s economy is largely agricultural, with coffee and tea as the main exports. Additionally, the region faces a shortage of skilled labour, particularly in the healthcare sector. Women who are employed in skilled labour can afford professional maternal care [20]. However, there is limited research on this topic in Burundi specifically.

In The Gambia, most of the women affected by poor pregnancy outcomes are not gainfully employed and mostly work in farms or as housewives with several burdens that kept them mostly physically and psychologically exhausted [50,60]. A study done in The Gambia, reported that the common occupations of women include farming, market vending, and domestic work. These occupations impact their pregnancies and births in several ways:

Physical demands: Farming and market vending involve physically demanding work that may require lifting heavy loads and working long hours. This can increase the risk of complications during pregnancy, such as premature labour, low birth weight, and fetal distress.

Limited access to healthcare: The women complained of their occupations giving them limited access to healthcare due to the nature of their work and financial constraints. This can result in delayed prenatal care and poor management of pregnancy-related conditions, such as anaemia, hypertension, and gestational diabetes.

Stress: Domestic work can be emotionally and physically stressful, which can have negative effects on maternal and fetal health. Stress during pregnancy has been linked to preterm labour, low birth weight, and developmental delays in children [60].

Occupation was found to have an impact on maternal and perinatal mortality rates in Nigeria [61]. Women working in agriculture, mining, and fishing industries had a higher risk of maternal mortality in Nigeria. The study suggested that occupational health and safety measures and improved access to maternal health services could help reduce maternal mortality rates among women in these industries [22] women in professional occupations were more likely to use maternal health services, such as antenatal care and skilled birth attendants, than women in other occupations in Nigeria [22,61].

Women who work for pay in Guinea had better access to ANC visits compared to those who did not, and this was because of financial empowerment [46]. Employment status is a predictor of wealth status in Guinea [27]. Also, majority of the women who were traders complained about been too occupied to attend antenatal clinics. Occupational status of the partners was also a factor to determine ANC attendance by their partners [48].

In Somalia it was established by that many of the mothers often have the obligation to support the family through hard and poorly paid jobs or by selling items in the market, even during pregnancy which may predispose them to stress which can lead to perinatal and maternal mortality [64]. Occupation was mentioned as one of the socioeconomic determinants of infant and maternal mortality in Chad [54].

## Discussion

The objective of this review is to elaborate on how specific socioeconomic factors (education, income and occupation) affect antenatal care service utilization as well as maternal and perinatal mortality in Sub-Saharan Africa. While the SES effect on the subject matter has been severely flogged, there remains limited evidence on how this SES specifically affects ANC and perinatal/maternal mortality. Hence, there is an urgent need to address this gap in the literature. Despite the global trend in policies of an almost-free to at-no-cost healthcare provisions for pregnant mothers, low level of income, poor and stressful occupation, and weak education has consistently been associated with poor birth outcomes [4,69]. This is particularly so in Sub-Saharan Africa as Nations in the West have better prognosis for fetal pregnancy outcomes, primarily attributed to the policies of care available to this population group in the form of health insurance.

The reviews reveal that the socio-economic factors limiting ANC utilization predispose women to high maternal and perinatal mortality and vary according to various regions in each country [59,70]. Categorizing these regions into two communities of rural and urban, overall, mortalities are higher in rural communities than in urban areas. Commonly in rural communities, there is a depleted healthcare system and the people are not able to transport to bigger cities for proper care, and this may invariably limit the ANC services provided, hence reducing utilization [26,71]. It is, therefore, essential that programs to empower and prompt better utilization of ANC should be focused on making it more accessible to the burdened communities [72].

Although the studies recorded other contributing SES factors across the countries’, education and income are the common SES affecting both ANC utilization and pregnancy outcome (perinatal and maternal mortality) [17,73,74]. Poor education of women and their spouses was peculiar to most of the findings in this study. The majority of the studies recommended that educating women will reduce the disparities in health-seeking behaviour which invariably improve ANC utilization. It is believed that improving education, particularly among women in SSA, will considerably minimize inequities in the consumption of ANC services and thereby reduce perinatal and maternal mortality.

Education has been identified ‘as a crucial factor in reducing both maternal and perinatal mortality rates across sub-Saharan Africa [6,39,75]. Women with no education had a significantly higher risk of maternal mortality than those with some. Increasing access to education for women, particularly in rural areas, can promote the use of ANC which could be an effective strategy for improving maternal and child health outcomes in the region [22,39].

Income was also revealed to be another limiting SES to the utilization of ANC due to limited access to appropriate medical treatment, which invariably exposes women to bad pregnancy outcomes (perinatal and maternal mortality) [71]. A significant inverse relationship between income and maternal mortality establishes that money is a considerable barrier to assessing comprehensive care in pregnancy and preventing birth complications to both mother and baby, many women may not be able to afford necessary care because of their personal limited income and that of their spouse [4,62,76]. A high risk of maternal death exists for those who cannot afford private hospitals and turn to mission homes and traditional means which further predisposes them to perinatal and maternal mortality [4,14,77].

Even though studies on the effect of occupation on ANC utilization and pregnancy outcome are limited. The review reveals that women’s occupation in countries with high mortality cases is burdening [78,79]. Even housewives who are believed or expected to be less busy, are more engaged physically and emotionally and are mostly health-challenged [59,80]. On the hand, women who are gainfully employed were revealed to have limited time for antenatal care services as well to report to the health facility early when some complications arise [26,81,82]. Therefore, it is essential to prioritize the health and safety of pregnant women in these occupations by providing access to adequate healthcare, reducing physical demands, and addressing environmental and occupational hazards. This can lead to improved pregnancy outcomes and better health for both the mother and child.

This study suggested that occupational status can impact maternal health outcomes in a country and that interventions to improve access to maternal health services should consider occupational differences, yet these have been poorly studied.

## Conclusion and Recommendation

In summary, the impact of poor socio-economic status on antenatal care services utilization and pregnancy outcomes in Sub-Saharan Africa is a significant public health issue. There is a clear link between low socio-economic status, low antenatal care utilization, and negative pregnancy outcomes, including maternal and perinatal mortality. Insufficient income and low levels of education among women in many countries in the region are the leading contributors to low antenatal care utilization and high perinatal and maternal mortality.

Efforts to improve the socio-economic status of women in Sub-Saharan Africa are essential to improving maternal and perinatal health outcomes. Initiatives such as increasing access to education, expanding access to healthcare services, and improving infrastructure in underserved areas can significantly impact. Additionally, community-based interventions that aim to improve awareness and knowledge about antenatal care, safe pregnancy, and childbirth practices can help to increase utilization and improve outcomes.

In conclusion, the impact of poor socio-economic status on antenatal care services utilization and pregnancy outcomes in Sub-Saharan Africa is a complex issue that requires a comprehensive and multidisciplinary approach. Improving the region’s maternal and perinatal health outcomes will require sustained efforts and collaboration between governments, healthcare providers, civil society organizations, and communities. Nonetheless, it is achievable, and the benefits will be immense, not only for mothers and newborns but also for the region’s overall development.

## Limitations and Future Directions

Occupational status and how it affects pregnancy outcomes as well as ANC visits was limited in all the studies identified. The impact of occupation is beyond the availability of money to pay for services related to healthcare, it also involves factors such as making time available to attend ANC and the nature of the job and how it can affect the outcome of pregnancy. Future studies must explore these factors, particularly through data collection in both quantitative and qualitative studies, to examine their impact.

One major limitation of this study is that most of the studies assessing the influence of socio-economic factors on pregnancy outcomes in SSA were done in rural communities where the problem was predominant. Education and income level were mostly implicated and thus recommended to empower women and educate them more, I would recommend for future research that similar studies should be done in peri-urban and urban cities where women are more empowered and educated to determine how these factors can also still negatively influence pregnancy outcomes and ANC visits.

There are variations to the socio-economic factors affecting pregnancy outcomes as well as antenatal care usage in Nigeria and need to be addressed differently, future studies should examine the role of community-level social influence on pregnancy outcomes particularly in the other regions aside from the northern part of the country [39].

## Data Availability

This scoping review does not involve the generation or analysis of original data. All data used in this review are derived from previously published studies, which are duly cited in the manuscript. As such, there are no additional datasets or raw data files associated with this review. All references to the included studies can be found in the reference section of the manuscript.

